# National trends in drug overdose mortality in Asian American, Native Hawaiian, and Pacific Islander populations, 2018-2022

**DOI:** 10.1101/2024.09.17.24313831

**Authors:** David T. Zhu, Andrew Park

## Abstract

**Background:** Drug overdose deaths have surged over the past two decades, disproportionately impacting racial/ethnic minority populations. Yet, little is known about drug overdose patterns among Asian American and Native Hawaiian/Pacific Islander (AANHPI) populations.

**Methods:** We obtained data on drug overdose deaths and population totals from the CDC WONDER Multiple Cause of Death database and American Community Survey between 2018 and 2022. We calculated crude mortality rates per 100,000, stratified by sex, US Census Division, and drug types—prescription opioids, heroin, fentanyl, cocaine, methamphetamine, and benzodiazepines. Additionally, we conducted disaggregated analyses for six Asian American subgroups (Asian Indian, Chinese, Filipino, Japanese, Korean, Vietnamese) and three NHPI subgroups (Hawaiian, Guamanian, Samoan).

**Results:** In 2022, there were 1226 drug overdose deaths among Asian Americans and 154 among NHPI individuals. The crude mortality rate for NHPI individuals (17.52 per 100,000; 95% CI: 14.76– 20.29) tripled that of Asian Americans (5.85 per 100,000; 95% CI: 5.52–6.18). Fentanyl was the leading cause of overdose deaths among Asian Americans (3.17 per 100,000; 95% CI: 2.93– 3.41), whereas methamphetamine was predominant among NHPI individuals (11.38 per 100,000; 95% CI: 9.15–13.61). Among Asian American subgroups, Japanese Americans had the highest mortality rate (9.90 per 100,000; 95% CI: 9.61–10.2), and among NHPI subgroups, Guamanians had the highest rates (43.16 per 100,000; 95% CI: 39.05–48.24).

**Conclusions:** These findings underscore the urgent need for culturally competent harm reduction services, mental health and addiction treatment, and social services, addressing structural barriers that perpetuate drug overdose disparities in AANHPI communities.

## INTRODUCTION

The drug overdose epidemic in the United States has dramatically risen over the past two decades, claiming more than 100,000 lives in 2023 (CDC, 2024) and representing a significant public health concern. Prior evidence demonstrates widening racial/ethnic disparities in drug overdose deaths in recent years, primarily attributed to the widespread proliferation of illicitly manufactured fentanyl, both alone and in combination with other substances in polysubstance formulations (Friedman & Shover, 2023; Zhu, 2024a). Notably, overdoses involving fentanyl mixed with stimulants (e.g., cocaine and methamphetamines) have contributed significantly to disproportionately high rates of drug overdose deaths among non-Hispanic Black and American Indian/Alaska Native (AIAN) populations (Friedman et al., 2022).

National surveys underscore the significant burden of substance use disorders among Asian American and Native Hawaiian/Pacific Islander (AANHPI) populations, estimating a prevalence of 1.5 million Asian American adults in 2020 (Choi et al., 2023). Nevertheless, research on substance use disorders and drug overdoses within AANHPI communities remains limited. A major contributing factor is the persistence of the ’model minority’ myth, which inaccurately portrays AANHPI individuals as uniformly successful and healthier across various conditions, including substance use disorders (Cheng et al., 2016; Walton & Truong, 2022).

This myth is further perpetuated by data systems that aggregate AANHPI individuals into a single, monolithic category, showing relatively low rates of drug overdose deaths compared to other racial/ethnic groups. However, these aggregated analyses obscure critical disparities across highly diverse AANHPI ethnic subgroups who possess unique social and cultural values, exhibit significant variations in physical and mental health burdens, and face multifaceted challenges in accessing healthcare (Subica et al., 2024; Zhu, 2024b). Well-documented variations in language barriers, health literacy, cultural perceptions of mental health and substance use, and access to evidence-based medications for opioid use disorder (OUD), such as methadone and buprenorphine, among other social determinations in health, likely contribute to the significant disparities in drug overdose deaths across AANHPI ethnic subgroups (Cheng et al., 2016; Walton & Truong, 2022; Choi et al., 2024; Ðoàn et al., 2024).

Given the paucity of research on drug overdoses in AANHPI populations, this study primarily aims to analyze national epidemiological trends in overdose deaths over time, geographical regions, and drug types. A secondary aim is to examine disparities across disaggregated AANHPI ethnic subgroups, offering insights to inform clinical and public health interventions that address the unique challenges faced by local AANHPI communities.

## METHODS

We obtained drug overdose mortality data from the CDC WONDER Multiple Cause of Death database for the years 2018 (the earliest year for which disaggregated data became available) to 2022. We defined drug overdoses using specific ICD-10 codes **(eMethods)**. For the aggregated Asian American and Native Hawaiian/Pacific Islander (NHPI) populations, we calculated crude mortality rates per 100,000 individuals. These rates were further stratified by sex, US Census Division, and specific drug types, including prescription opioids, heroin, fentanyl, cocaine, methamphetamine, and benzodiazepines.

Additionally, we performed disaggregated analyses across six Asian American ethnic subgroups (Asian Indian, Chinese, Filipino, Japanese, Korean, and Vietnamese) and three NHPI subgroups (Hawaiian, Guamanian, and Samoan). Individuals not classified into these categories were categorized as ’Other Asians’ or ’Other Pacific Islanders.’ While overdose mortality data were sourced from CDC WONDER, the database does not provide population totals for disaggregated subgroups. Thus, we obtained population estimates from the American Community Survey (ACS) for each ethnic subgroup and year, following established methodologies (Bui et al., 2024). Crude mortality rates per 100,000 were calculated using overdose deaths from CDC WONDER (numerator) and ACS population totals (denominator).

Data analysis and visualization were conducted using R software (version 4.0.3). We adhered to the STROBE guidelines for cross-sectional studies. The Virginia Commonwealth University Institutional Review Board deemed this study exempt from review, as it did not involve human subjects research.

## RESULTS

In 2022, there were 1226 drug overdoses among Asian Americans and 154 drug overdoses among NHPI individuals. The crude mortality rate was markedly higher for NHPI individuals, at 17.52 per 100,000 (95% CI: 14.76–20.29), compared to 5.85 per 100,000 (95% CI: 5.52–6.18) among Asian Americans **(Table 1)**. Between 2018 and 2022, drug overdose death rates increased by 75% among Asian Americans and 65% among NHPI individuals.

**Table 1.**
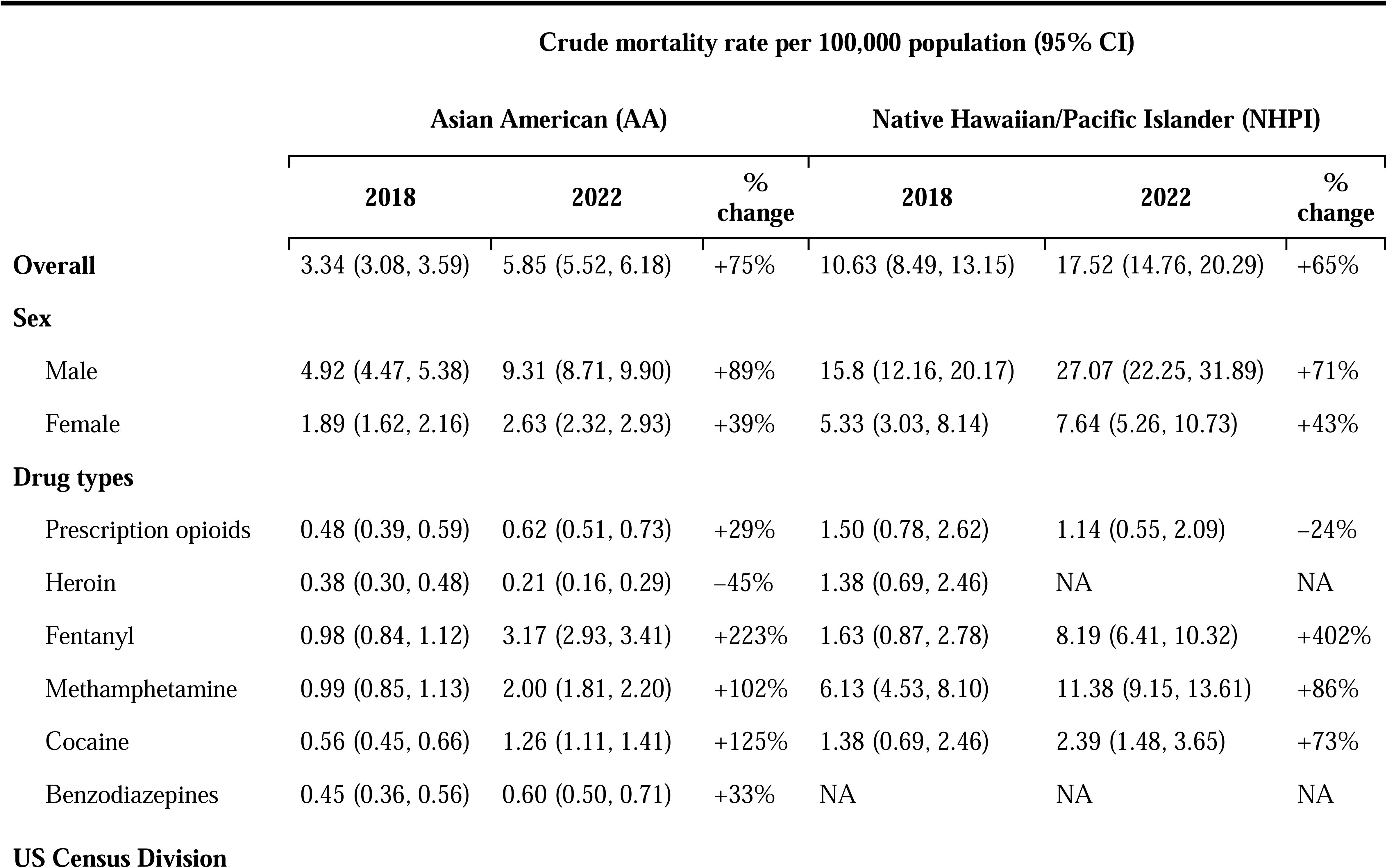
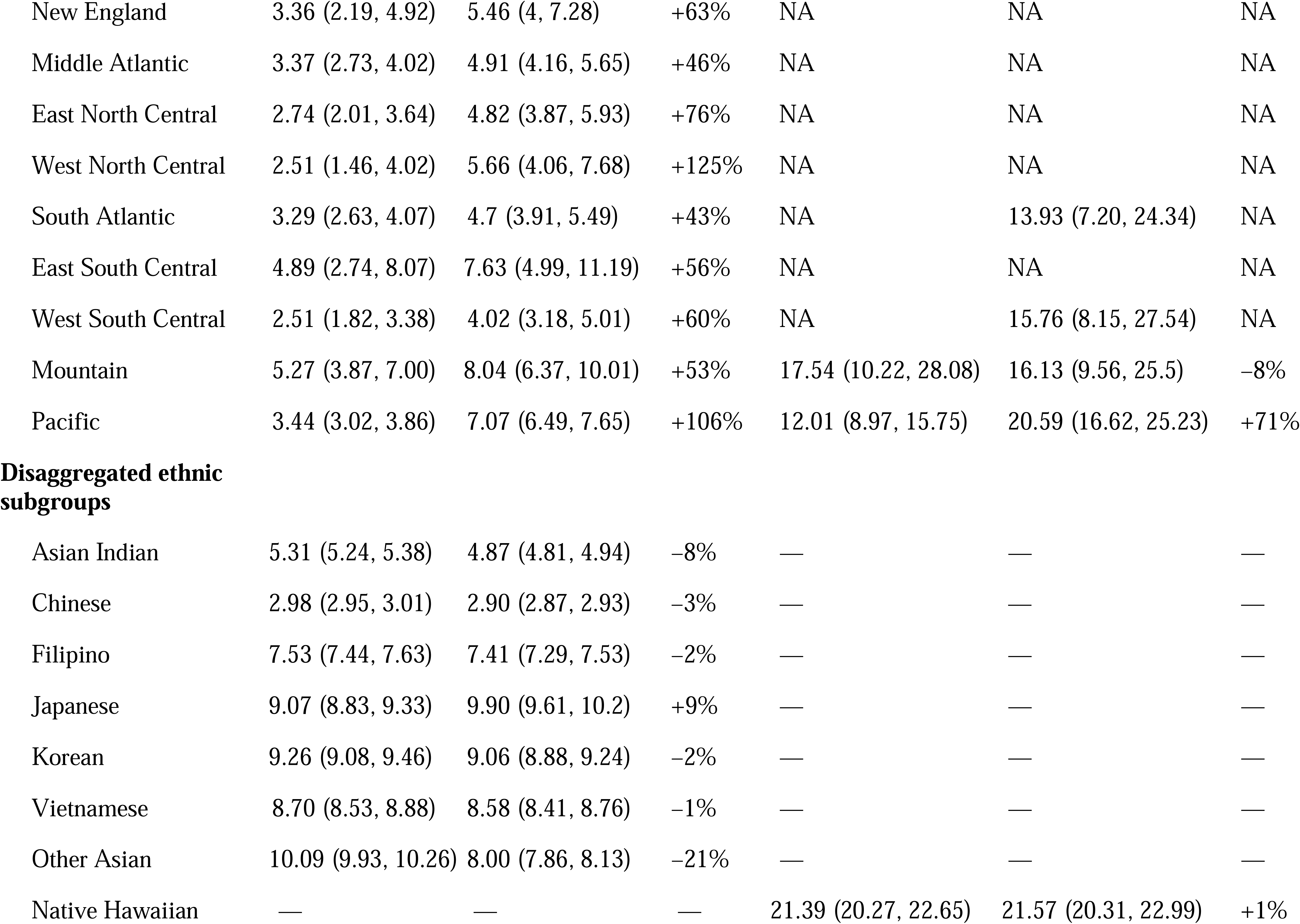
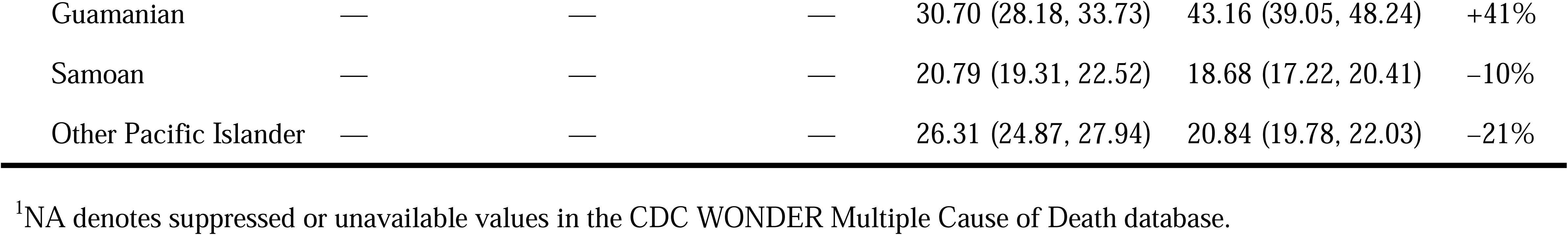
Drug overdose crude mortality rates per 100,000 among Asian American and Native Hawaiian/Pacific Islander populations, 2018 to 2022^1^.

| Crude mortality rate per 100,000 population (95% CI) |  |  |  |  |  |  |
| --- | --- | --- | --- | --- | --- | --- |
|  | Asian American (AA) |  |  | Native Hawaiian/Pacific Islander (NHPI) |  |  |
|  | 2018 | 2022 | % change | 2018 | 2022 | % change |
| **Overall** | 3.34 (3.08, 3.59) | 5.85 (5.52, 6.18) | +75% | 10.63 (8.49, 13.15) | 17.52 (14.76, 20.29) | +65% |
| **Sex** |  |  |  |  |  |  |
| Male | 4.92 (4.47, 5.38) | 9.31 (8.71, 9.90) | +89% | 15.8 (12.16, 20.17) | 27.07 (22.25, 31.89) | +71% |
| Female | 1.89 (1.62, 2.16) | 2.63 (2.32, 2.93) | +39% | 5.33 (3.03, 8.14) | 7.64 (5.26, 10.73) | +43% |
| **Drug types** |  |  |  |  |  |  |
| Prescription opioids | 0.48 (0.39, 0.59) | 0.62 (0.51, 0.73) | +29% | 1.50 (0.78, 2.62) | 1.14 (0.55, 2.09) | −24% |
| Heroin | 0.38 (0.30, 0.48) | 0.21 (0.16, 0.29) | −45% | 1.38 (0.69, 2.46) | NA | NA |
| Fentanyl | 0.98 (0.84, 1.12) | 3.17 (2.93, 3.41) | +223% | 1.63 (0.87, 2.78) | 8.19 (6.41, 10.32) | +402% |
| Methamphetamine | 0.99 (0.85, 1.13) | 2.00 (1.81, 2.20) | +102% | 6.13 (4.53, 8.10) | 11.38 (9.15, 13.61) | +86% |
| Cocaine | 0.56 (0.45, 0.66) | 1.26 (1.11, 1.41) | +125% | 1.38 (0.69, 2.46) | 2.39 (1.48, 3.65) | +73% |
| Benzodiazepines | 0.45 (0.36, 0.56) | 0.60 (0.50, 0.71) | +33% | NA | NA | NA |
| **US Census Division** |  |  |  |  |  |  |
| New England | 3.36 (2.19, 4.92) | 5.46 (4, 7.28) | +63% | NA | NA | NA |
| Middle Atlantic | 3.37 (2.73, 4.02) | 4.91 (4.16, 5.65) | +46% | NA | NA | NA |
| East North Central | 2.74 (2.01, 3.64) | 4.82 (3.87, 5.93) | +76% | NA | NA | NA |
| West North Central | 2.51 (1.46, 4.02) | 5.66 (4.06, 7.68) | +125% | NA | NA | NA |
| South Atlantic | 3.29 (2.63, 4.07) | 4.7 (3.91, 5.49) | +43% | NA | 13.93 (7.20, 24.34) | NA |
| East South Central | 4.89 (2.74, 8.07) | 7.63 (4.99, 11.19) | +56% | NA | NA | NA |
| West South Central | 2.51 (1.82, 3.38) | 4.02 (3.18, 5.01) | +60% | NA | 15.76 (8.15, 27.54) | NA |
| Mountain | 5.27 (3.87, 7.00) | 8.04 (6.37, 10.01) | +53% | 17.54 (10.22, 28.08) | 16.13 (9.56, 25.5) | −8% |
| Pacific | 3.44 (3.02, 3.86) | 7.07 (6.49, 7.65) | +106% | 12.01 (8.97, 15.75) | 20.59 (16.62, 25.23) | +71% |
| **Disaggregated ethnic subgroups** |  |  |  |  |  |  |
| Asian Indian | 5.31 (5.24, 5.38) | 4.87 (4.81, 4.94) | −8% | — | — | — |
| Chinese | 2.98 (2.95, 3.01) | 2.90 (2.87, 2.93) | −3% | — | — | — |
| Filipino | 7.53 (7.44, 7.63) | 7.41 (7.29, 7.53) | −2% | — | — | — |
| Japanese | 9.07 (8.83, 9.33) | 9.90 (9.61, 10.2) | +9% | — | — | — |
| Korean | 9.26 (9.08, 9.46) | 9.06 (8.88, 9.24) | −2% | — | — | — |
| Vietnamese | 8.70 (8.53, 8.88) | 8.58 (8.41, 8.76) | −1% | — | — | — |
| Other Asian | 10.09 (9.93, 10.26) | 8.00 (7.86, 8.13) | −21% | — | — | — |
| Native Hawaiian | — | — | — | 21.39 (20.27, 22.65) | 21.57 (20.31, 22.99) | +1% |
| Guamanian | — | — | — | 30.70 (28.18, 33.73) | 43.16 (39.05, 48.24) | +41% |
| Samoan | — | — | — | 20.79 (19.31, 22.52) | 18.68 (17.22, 20.41) | –10% |
| Other Pacific Islander | — | — | — | 26.31 (24.87, 27.94) | 20.84 (19.78, 22.03) | –21% |

<sup>1</sup>NA denotes suppressed or unavailable values in the CDC WONDER Multiple Cause of Death database.

Sex disparities were notable **(Table 1)**. In 2022, Asian American males experienced overdose death rates 3.54 times higher (9.31 per 100,000; 95% CI: 8.71–9.90) than their female counterparts (2.63 per 100,000; 95% CI: 2.32–2.93). Similarly, NHPI males had overdose rates 3.54 times higher (27.07 per 100,000; 95% CI: 22.25–31.89) compared to NHPI females (7.64 per 100,000; 95% CI: 5.26–10.73).

Disparities across drug types were also pronounced **(Table 1)**. In 2022, fentanyl was the leading cause of overdose deaths among Asian Americans (3.17 per 100,000; 95% CI: 2.93–3.41) and the second leading cause among NHPI individuals (8.19 per 100,000; 95% CI: 6.41–10.32), with NHPI rates 2.58 times higher. Conversely, methamphetamine was the leading cause of overdose deaths among NHPI individuals (11.38 per 100,000; 95% CI: 9.15–13.61) and the second leading cause among Asian Americans (2.00 per 100,000; 95% CI: 1.81–2.20), with NHPI rates 5.69 times higher. Cocaine-related overdoses were the third leading cause of overdose deaths among both groups, with Asian Americans at 1.26 per 100,000 (95% CI: 1.11–1.41) and NHPI individuals at 2.39 per 100,000 (95% CI: 1.48–3.65).

Between 2018 and 2022, fentanyl overdoses surged by 223% among Asian Americans and 402% among NHPI individuals **(Table 1)**. Methamphetamine and cocaine involvement showed similar increases among both Asian Americans (by 102% and 125%, respectively) and NHPI individuals (by 86% and 73%, respectively).

Geographical variations were significant **(Table 1)**. In 2022, Asian Americans had the highest overdose death rates in the Mountain (8.04 per 100,000; 95% CI: 6.37–10.01) and Pacific (7.07 per 100,000; 95% CI: 6.49–7.65) US Census Divisions. For NHPI individuals, the Pacific Division had the highest rate (20.59 per 100,000; 95% CI: 16.62–25.23), followed by the Mountain Division (16.13 per 100,000; 95% CI: 9.56–25.5). From 2018 to 2022, the West North Central Division saw the greatest increase (125%) among Asian Americans, while the Pacific Division experienced the highest increase (71%) among NHPI individuals.

Additionally, disaggregation of AANHPI ethnic subgroups revealed pronounced disparities in overdose mortality rates **(Table 1**, **Figure 1, and eTable 1 in the Supplement)**. In 2022, Japanese Americans had the highest overdose mortality rate (9.90 per 100,000; 95% CI: 9.61– 10.2), more than triple the rate among Chinese Americans, who had the lowest rate (2.90 per 100,000; 95% CI: 2.87–2.93). Among NHPI subgroups, Guamanians showed the highest rates (43.16 per 100,000; 95% CI: 39.05–48.24), more than double the rate among Samoans, who had the lowest rate (18.68 per 100,000; 95% CI: 17.22–20.41).

**Figure 1.**
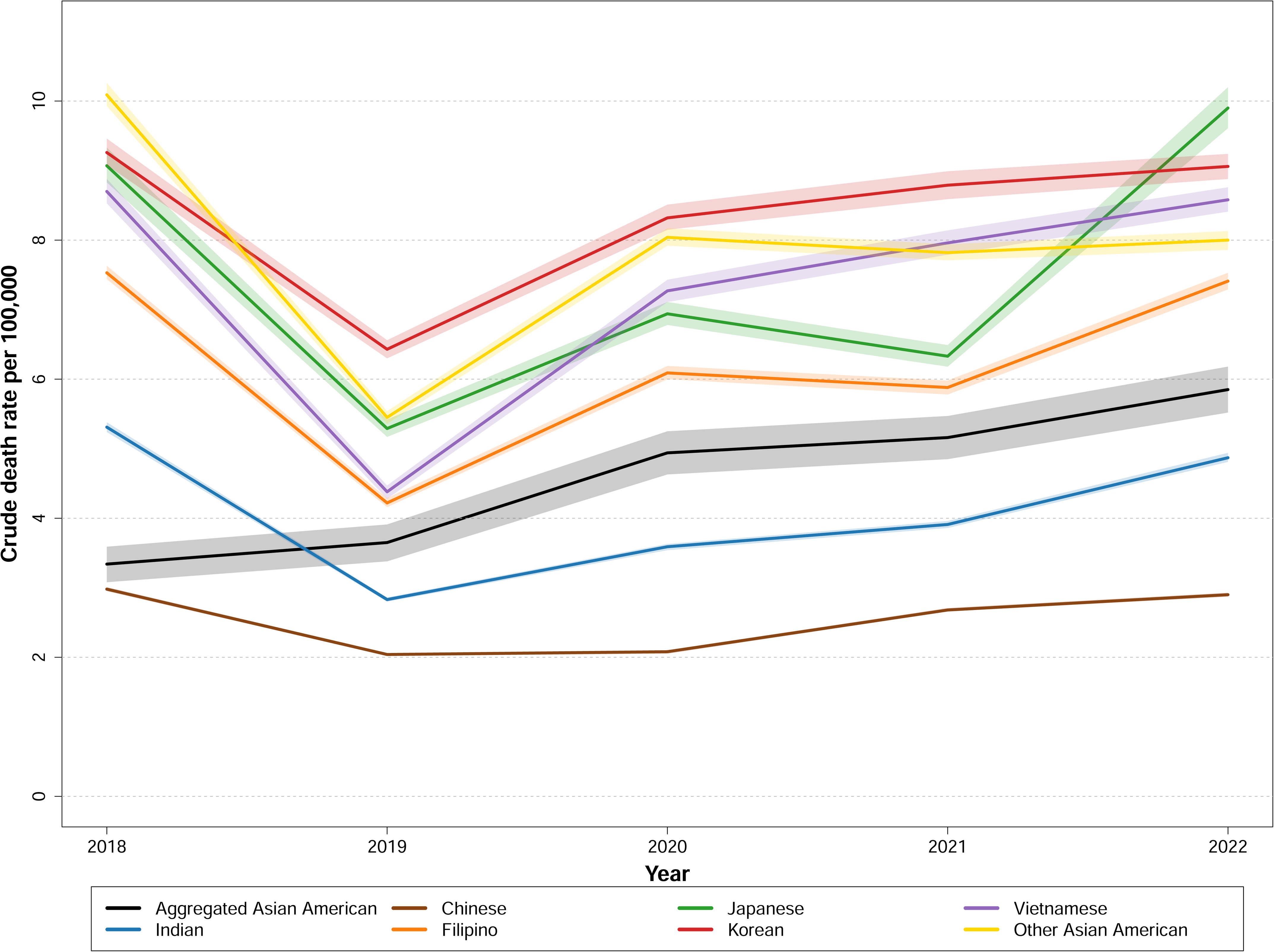
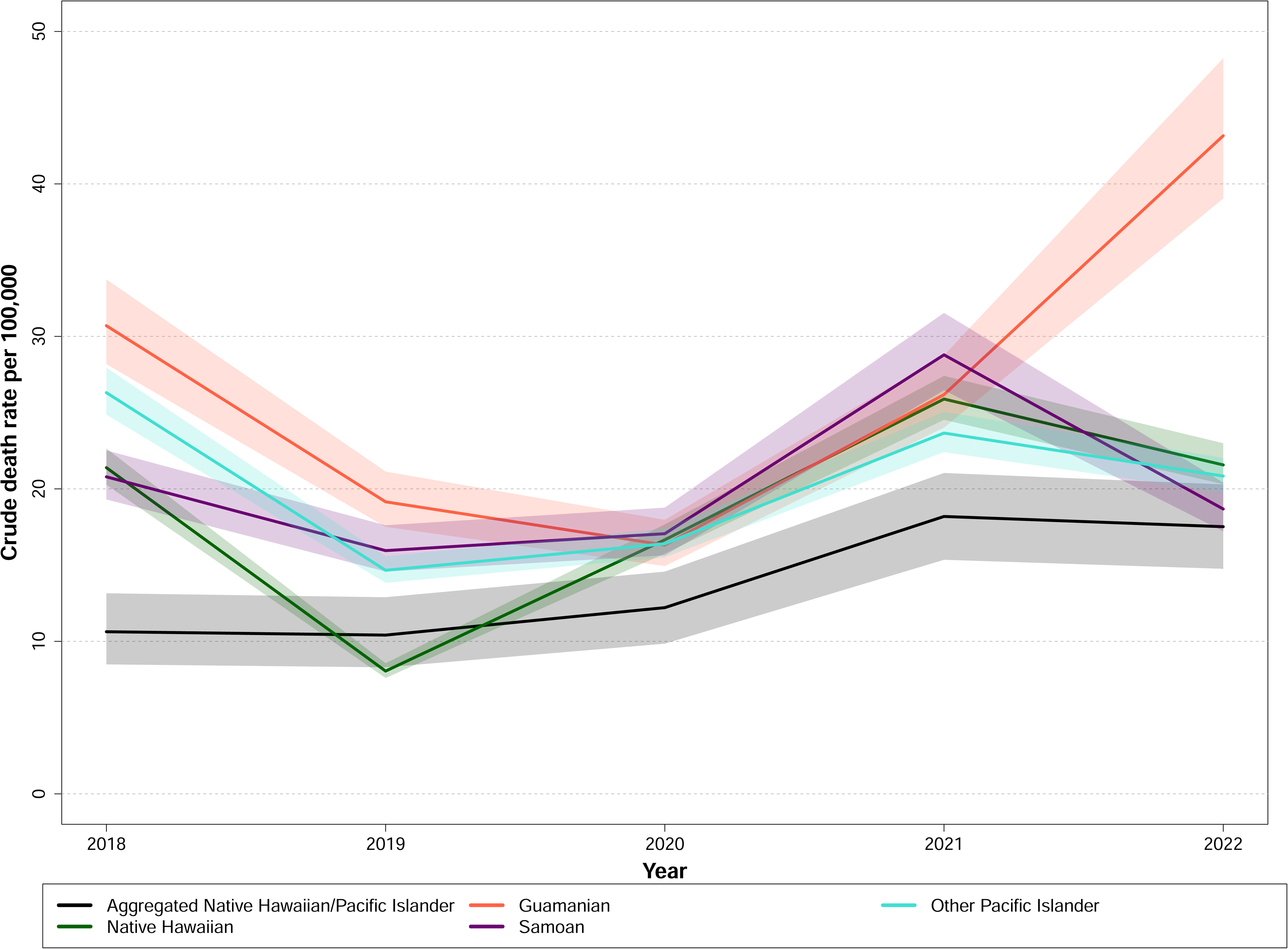
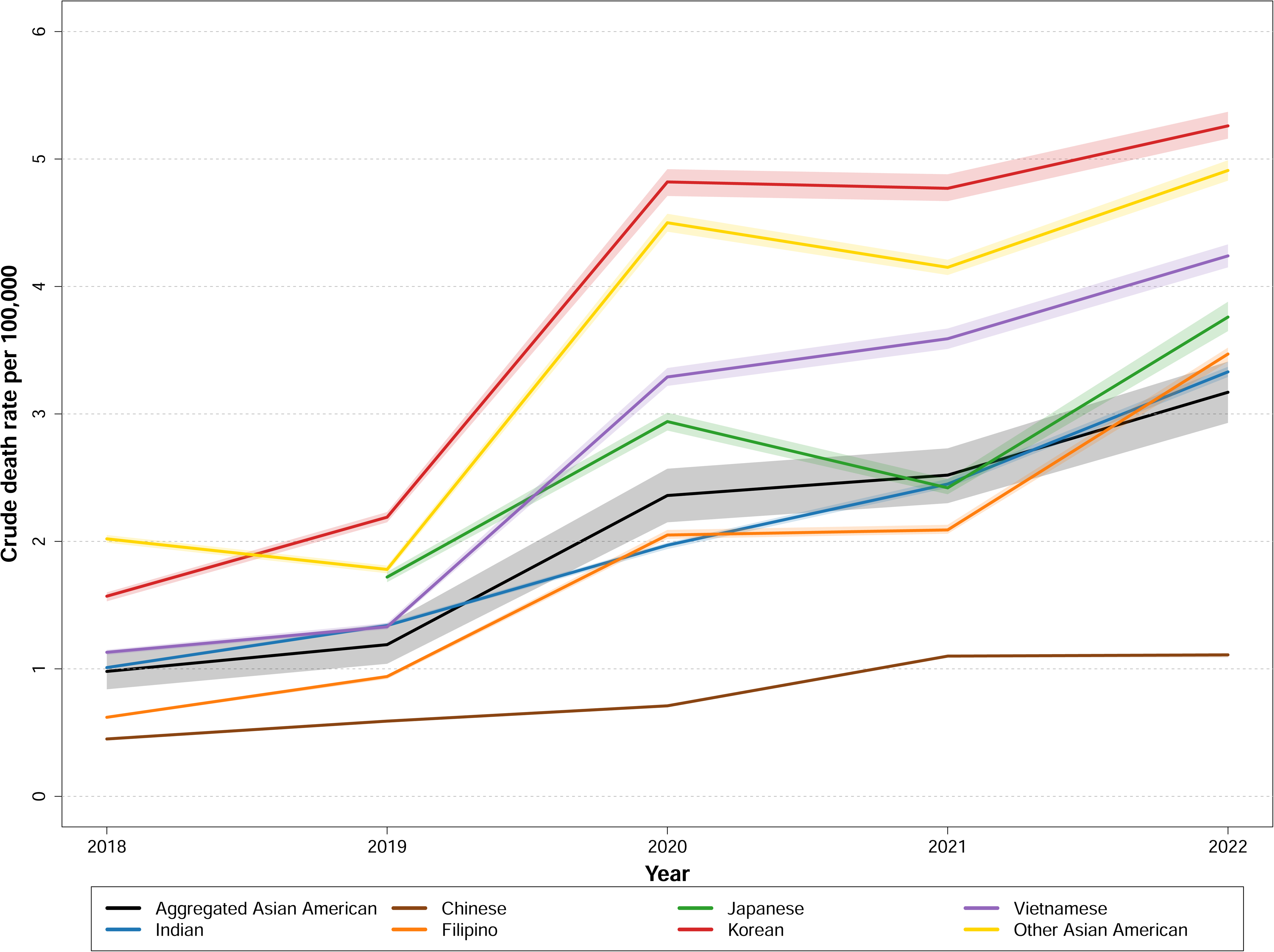
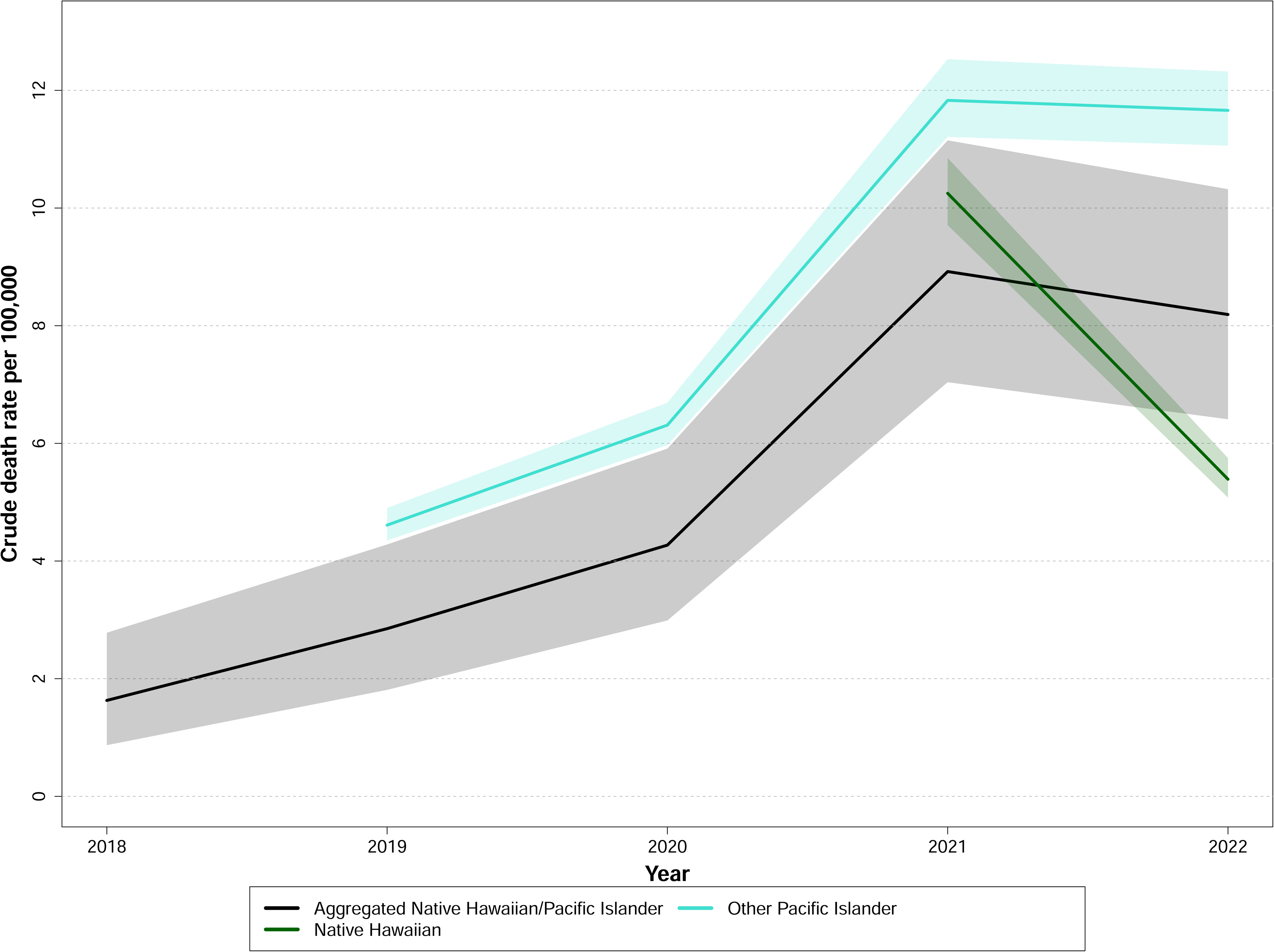
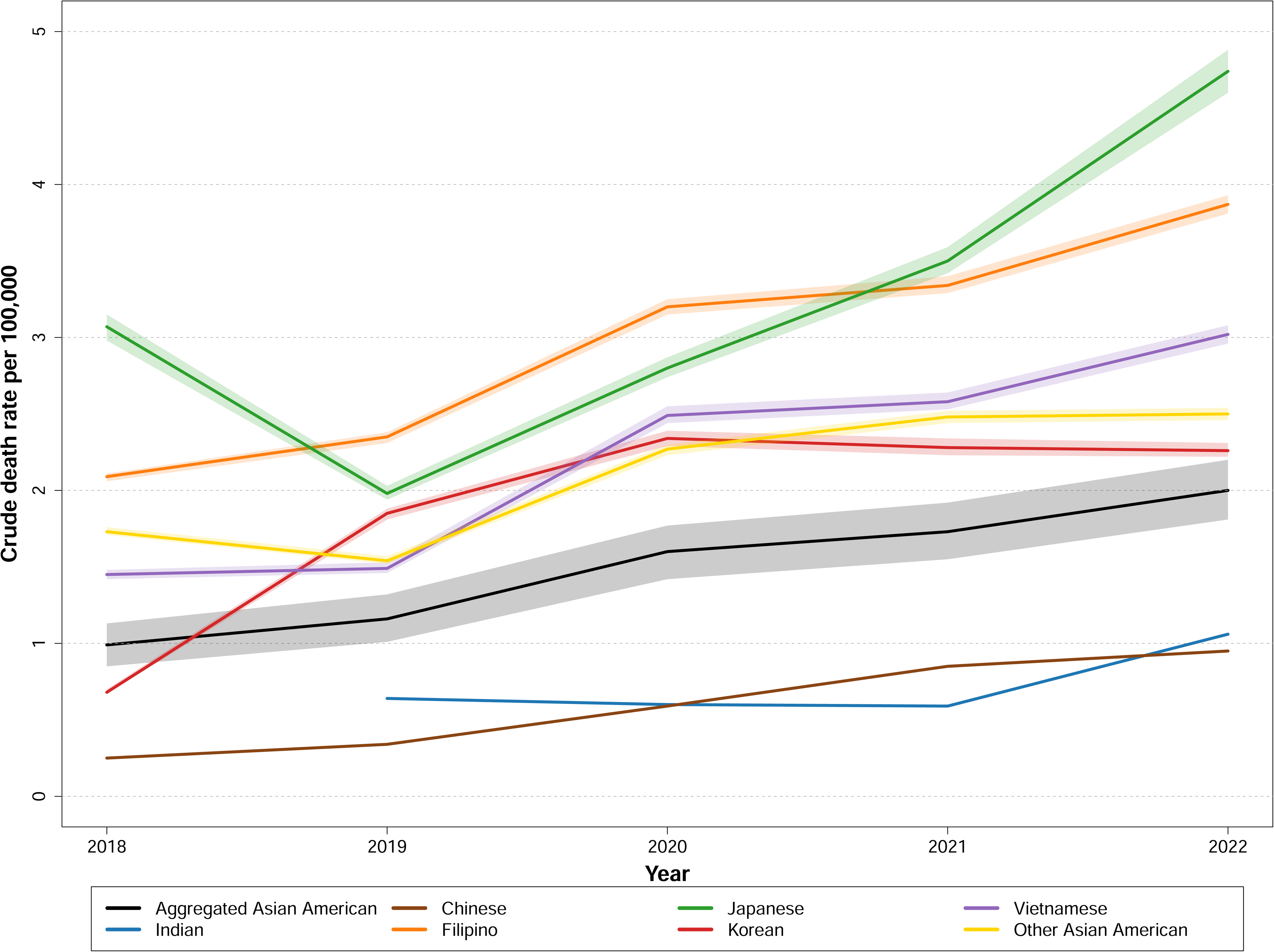
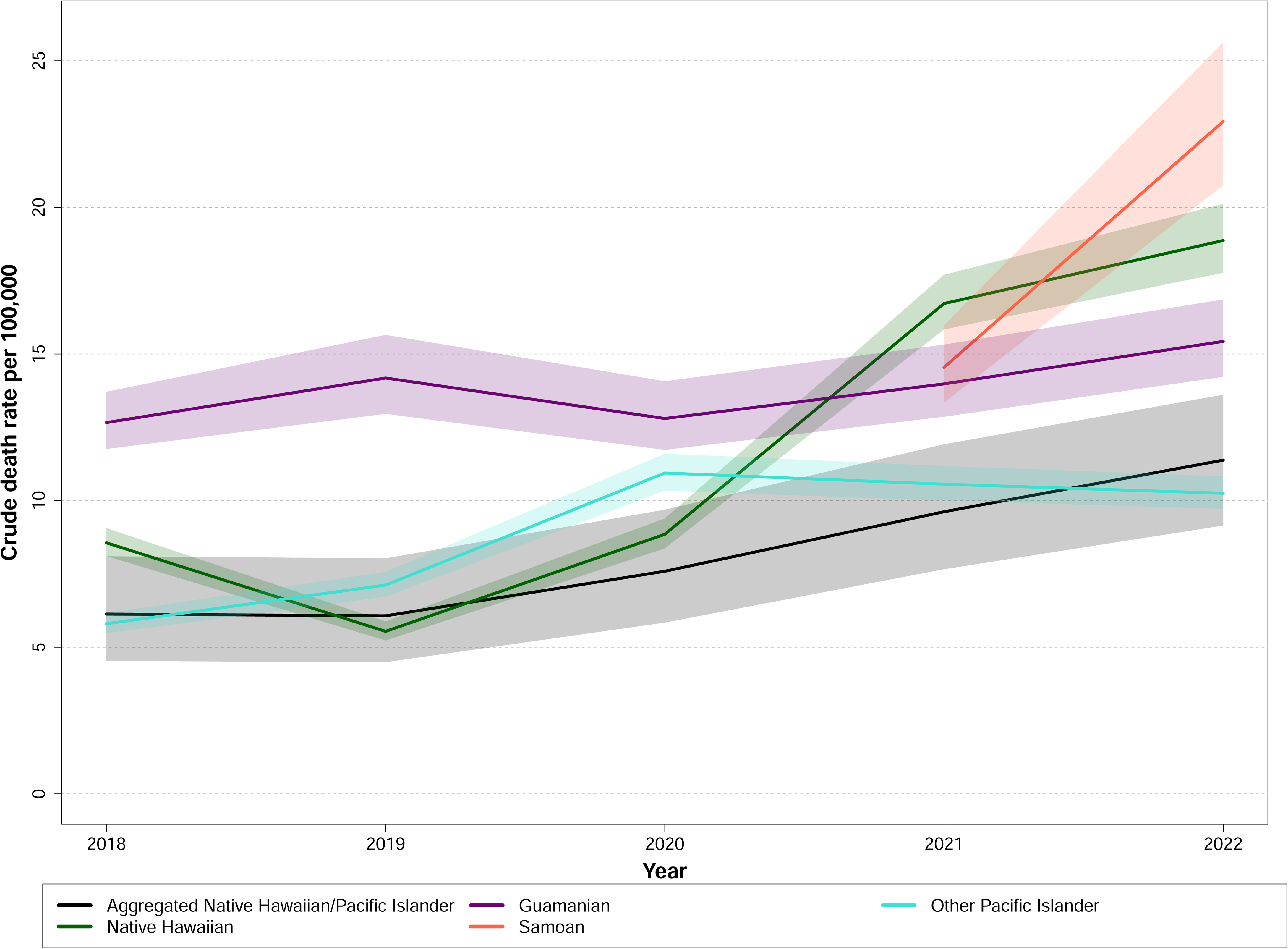
Crude mortality rates for drug overdoses among aggregated and disaggregated Asian American and Native Hawaiian/Pacific Islander ethnic groups, 2018-2022. Crude mortality rates (per 100,000) from 2018-2022 are presented for disaggregated Asian American (AA) subgroups in the following categories: **(A)** overall drug overdoses, (**C**) fentanyl overdoses, and **(E)** methamphetamine overdoses. Six AA ethnic subgroups were included: Asian Indian, Chinese, Filipino, Japanese, Korean, and Vietnamese. Similarly, crude mortality rates (per 100,000) from 2018-2022 were also presented for disaggregated Native Hawaiian/Pacific Islander (NHPI) subgroups in (B) overall drug overdoses, (D) fentanyl overdoses, and (F) methamphetamine overdoses. The NHPI ethnic subgroups included Hawaiian, Guamanian, and Samoan individuals. Shaded regions denote 95% confidence intervals (CIs). Subgroups without any consecutive yearly data were excluded from the figure. Data were sourced from the CDC WONDER Multiple Cause of Death database. A. Total drug overdoses, Asian Americans B. Total drug overdoses, Native Hawaiians/Pacific Islanders C. Fentanyl overdoses, Asian Americans D. Fentanyl overdoses, Native Hawaiians/Pacific Islanders E. Methamphetamine overdoses, Asian Americans F. Methamphetamine overdoses, Native Hawaiians/Pacific Islanders

Regarding specific drug types, Korean Americans had the highest fentanyl overdose rates in 2022 (5.26 per 100,000; 95% CI: 5.16–5.37), a 235% increase since 2018 **(Figure 1 and eTables 2-3 in the Supplement)**. Japanese Americans had the highest methamphetamine overdose rates (4.74 per 100,000; 95% CI: 4.60–4.88), reflecting a 54% increase since 2018. Guamanians had the highest rates for both fentanyl (26.98 per 100,000; 95% CI: 24.41, 30.15) and methamphetamine (22.93 per 100,000; 95% CI: 20.75–25.63) overdose deaths, with methamphetamine-related deaths increasing by 60% since 2021 (data from 2018-2020 were unavailable).

## DISCUSSION

Recent studies have increasingly highlighted racial and ethnic disparities in drug overdose mortality, yet AANHPI populations have often been underrecognized in this discourse. Our findings indicate a concerning rise in overdose death rates among both Asian Americans and NHPI populations from 2018 to 2022, with the NHPI rate tripling that of Asian Americans by 2022. These disparities were particularly pronounced among males and those living in the Mountain and Pacific Census Divisions. Further, disaggregated data reveals substantial variations across AANHPI ethnic subgroups, with Guamanians exhibiting higher rates — double that of the aggregated NHPI population. These trends underscore the urgent need for culturally tailored interventions that address the structural inequities perpetuating these inequities, alongside more detailed and granular epidemiological data collection on AANHPI ethnic subgroups to identify and address critical disparities.

The disproportionate burden of drug overdose deaths among NHPI populations must be considered in light of the underlying social and structural determinants of health, including socioeconomic status, housing stability, health literacy, language barriers, and access to evidence-based addiction treatments and medications like methadone and buprenorphine (Choi et al., 2023; Zhu et al., 2024b). Prior evidence suggests that, compared to Asian Americans, NHPI individuals are significantly more likely to be unemployed, have less than a high school education, live under the federal poverty line, and face financial barriers to treatment (Morisako et al., 2017). In Hawaii, Native Hawaiians, who comprise only 13% of the population, represent around 70% of the homeless population — with 80% of these homeless individuals testing positive for methamphetamine and fentanyl use (Subica et al., 2024).

The high prevalence of other substance use disorders and mental health conditions among NHPI populations, such as alcohol use disorder, anxiety, and depression, further heightens the risk of substance use disorders and drug overdoses (Wyatt et al., 2015; Burrage et al., 2020; Choi et al., 2024). Moreover, the surge in anti-AANHPI sentiment and discrimination during the COVID-19 pandemic has intensified the mental health burden in these populations, potentially driving greater substance use as a coping mechanism (Choi et al., 2024). Additionally, prior evidence shows that substance use treatment utilization among Asian Americans is low, with only 3% seeking treatment (Von Gunten & Wu, 2021), while three out of five NHPI individuals in need of treatment either avoided or delayed it (Choi et al., 2024). Another study demonstrates that AANHPI populations have lower rates of receiving buprenorphine compared to their White counterparts (Dunphy et al., 2022).

These trends underscore the critical intersections between substance use and structural inequities within AANHPI communities, highlighting the urgent need to enhance culturally tailored mental health and addiction services. Further efforts are also needed to provide AANHPI communities with broader access to health education regarding evidence-based and destigmatizing information about substance use disorders, and expanding the accessibility of primary care and addiction specialists to address local treatment needs. Additionally, improving data systems and epidemiological surveillance, particularly by incorporating disaggregated AANHPI data that reflects the diverse ethnic subgroups within this population, is crucial for implementing targeted clinical and public health interventions to address disparities across AANHPI ethnic subgroups. For instance, our findings reveal a disproportionate increase in drug overdose death rates among Guamanians, primarily attributed to methamphetamine overdoses, which is consistent with increased methamphetamine availability in Guam (DEA, 2020). This underscores the urgent need for culturally competent harm reduction services tailored to local needs among Guamanian communities, including substance use counseling, and overdose prevention education, while addressing structural inequities such as poverty, housing instability, among many others.

### Limitations

This study faces several limitations. It relies on CDC WONDER death certificate data, prone to racial misclassification of AANHPI subgroups (Holland & Palaniappan, 2015). Further, the cross-sectional design precludes causal inferences, thus, future prospective or quasi-experimental studies are warranted. Moreover, the study was constrained by the limited range of AANHPI subgroups defined in the CDC WONDER database, which does not fully capture the vast diversity within AANHPI populations. For instance, while the American Community Survey (ACS) recognizes 21 distinct Asian American subgroups (Monte & Shin, 2022), our analysis was restricted to only six subgroups available in CDC WONDER. Finally, future research should also explore social determinants — including socioeconomic status, health literacy, immigration status, and healthcare access — that could contribute to disparities in drug overdose deaths in AANHPI populations.

## CONCLUSION

This study underscores rising drug overdose mortality rates in AANHPI populations, with notable disparities across sociodemographic characteristics and geographical regions. Targeted clinical and public health interventions are urgent needed to improve access to culturally competent addiction treatment, mental health resources, and social services. Addressing these needs is crucial for overcoming structural barriers in drug overdose prevention and treatment, aiming to promote health equity in AANHPI populations.

## Author disclosures

### CRediT authorship contribution statement

David T. Zhu: Conceptualization, data curation, formal analysis, methodology, supervision, writing – original draft, writing – reviewing & editing. Andrew Park: Conceptualization, writing – original draft, writing – reviewing & editing.

### Data statement

The data that support the findings of this study are publicly available at https://wonder.cdc.gov/mcd.html and https://www.census.gov/programs-surveys/acs/data.html

## Supporting information

eMethods

## Data Availability

https://www.census.gov/programs-surveys/acs/data.html

