## Supplementary material for "National trends in drug overdose mortality in Asian American, Native Hawaiian, and Pacific Islander populations, 2018-2022": eMethods

We obtained data from death certificates in the publicly available Centers for Disease Control and Prevention Wide-Ranging Online Data for Epidemiologic Research (CDC WONDER) Multiple Cause of Death database (<https://wonder.cdc.gov/mcd.html>).

We defined deaths associated with ‘drug overdose’ using the International Classification of Diseases, 10th revision (ICD-10) codes X40-44 (unintentional drug poisonings), X60-64 (intentional drug poisonings), X85 (drug poisoning assaults), and Y10-Y14 (drug poisonings of undetermined intent) as underlying causes of death, consistent with established methodologies [(Friedman et al., 2022)](https://psychiatryonline.org/doi/10.1176/appi.ajp.2021.21040381).

Specific drug types were identified using the following ICD-10 codes: T40.1 (heroin), T40.2/40.3 (prescription opioids), T40.4 (fentanyls and synthetic analogs), T40.5 (cocaine), T42.4 (benzodiazepines), and T43.6 (methamphetamine) as multiple causes of death, consistent with previous studies [(Friedman et al., 2022)](https://psychiatryonline.org/doi/10.1176/appi.ajp.2021.21040381).
