## Supplement for "National trends in drug overdose mortality in Asian American, Native Hawaiian, and Pacific Islander populations, 2018-2022"

**eTable 1. Drug overdose crude mortality rates per 100,000 among disaggregated Asian American and Native Hawaiian/Pacific Islander populations, 2018 to 2022**

|  | **Crude mortality rate per 100,000 population (95% CI)** | | | | |
| --- | --- | --- | --- | --- | --- |
| **Race** | **2018** | **2019** | **2020** | **2021** | **2022** |
| Asian Americans (aggregated) | 3.34 (3.08, 3.59) | 3.65 (3.38, 3.91) | 4.94 (4.63, 5.25) | 5.16 (4.85, 5.47) | 5.85 (5.52, 6.18) |
| Asian Indian | 5.31 (5.24, 5.38) | 2.83 (2.80, 2.86) | 3.59 (3.54, 3.63) | 3.91 (3.86, 3.96) | 4.87 (4.81, 4.94) |
| Chinese | 2.98 (2.95, 3.01) | 2.04 (2.02, 2.06) | 2.08 (2.05, 2.10) | 2.68 (2.66, 2.71) | 2.9 (2.87, 2.93) |
| Filipino | 7.53 (7.44, 7.63) | 4.22 (4.16, 4.29) | 6.09 (6.00, 6.19) | 5.88 (5.78, 5.98) | 7.41 (7.29, 7.53) |
| Japanese | 9.07 (8.83, 9.33) | 5.29 (5.17, 5.42) | 6.94 (6.78, 7.11) | 6.33 (6.18, 6.49) | 9.9 (9.61, 10.20) |
| Korean | 9.26 (9.08, 9.46) | 6.43 (6.30, 6.56) | 8.32 (8.15, 8.51) | 8.79 (8.59, 8.99) | 9.06 (8.88, 9.24) |
| Vietnamese | 8.7 (8.53, 8.88) | 4.38 (4.28, 4.47) | 7.27 (7.11, 7.43) | 7.96 (7.79, 8.14) | 8.58 (8.41, 8.76) |
| Other Asian | 10.09 (9.93, 10.26) | 5.45 (5.36, 5.54) | 8.04 (7.92, 8.17) | 7.82 (7.71, 7.94) | 8 (7.86, 8.13) |
| Native Hawaiians/Pacific Islanders (aggregated) | 10.63 (8.49, 13.15) | 10.41 (8.30, 12.89) | 12.21 (9.85, 14.57) | 18.19 (15.35, 21.04) | 17.52 (14.76, 20.29) |
| Hawaiian | 21.39 (20.27, 22.65) | 8.05 (7.60, 8.56) | 16.66 (15.76, 17.67) | 25.89 (24.53, 27.41) | 53.95 (48.82, 60.3) |
| Guamanian | 30.70 (28.18, 33.73) | 19.15 (17.51, 21.13) | 16.32 (14.94, 17.97) | 26.18 (24.02, 28.76) | 17.25 (16.25, 18.39) |
| Samoan | 20.79 (19.31, 22.52) | 15.95 (14.58, 17.61) | 17.06 (15.64, 18.77) | 28.79 (26.47, 31.54) | 18.68 (17.22, 20.41) |
| Other Pacific Islander | 26.31 (24.87, 27.94) | 14.66 (13.84, 15.58) | 16.40 (15.51, 17.4) | 23.66 (22.41, 25.05) | 20.84 (19.78, 22.03) |

**eTable 2. Fentanyl overdose crude mortality rates per 100,000 among disaggregated Asian American and Native Hawaiian/Pacific Islander populations, 2018 to 2022**

|  | **Crude mortality rate per 100,000 population (95% CI)** | | | | |
| --- | --- | --- | --- | --- | --- |
| **Race** | **2018** | **2019** | **2020** | **2021** | **2022** |
| Asian Americans (aggregated) | 0.98 (0.84, 1.12) | 1.19 (1.04, 1.35) | 2.36 (2.15, 2.57) | 2.52 (2.30, 2.73) | 3.17 (2.93, 3.41) |
| Asian Indian | 1.01 (1.00, 1.02) | 1.34 (1.33, 1.36) | 1.97 (1.94, 1.99) | 2.45 (2.42, 2.49) | 3.33 (3.29, 3.37) |
| Chinese | 0.45 (0.45, 0.46) | 0.59 (0.58, 0.6) | 0.71 (0.70, 0.71) | 1.10 (1.09, 1.11) | 1.11 (1.09, 1.12) |
| Filipino | 0.62 (0.61, 0.62) | 0.94 (0.92, 0.95) | 2.05 (2.02, 2.09) | 2.09 (2.06, 2.13) | 3.47 (3.41, 3.52) |
| Japanese | NA | 1.72 (1.68, 1.76) | 2.94 (2.87, 3.01) | 2.42 (2.37, 2.49) | 3.76 (3.65, 3.88) |
| Korean | 1.57 (1.53, 1.60) | 2.19 (2.15, 2.23) | 4.82 (4.71, 4.92) | 4.77 (4.67, 4.88) | 5.26 (5.16, 5.37) |
| Vietnamese | 1.13 (1.11, 1.15) | 1.33 (1.31, 1.36) | 3.29 (3.22, 3.36) | 3.59 (3.51, 3.67) | 4.24 (4.15, 4.33) |
| Other Asian | 2.02 (1.99, 2.05) | 1.78 (1.75, 1.81) | 4.5 (4.43, 4.57) | 4.15 (4.09, 4.21) | 4.91 (4.83, 4.99) |
| Native Hawaiians/Pacific Islanders (aggregated) | 1.63 (0.87, 2.78) | 2.85 (1.81, 4.28) | 4.27 (2.99, 5.91) | 8.92 (7.04, 11.15) | 8.19 (6.41, 10.32) |
| Hawaiian | NA | NA | NA | 10.25 (9.71, 10.85) | 5.39 (5.08, 5.75) |
| Guamanian | NA | NA | NA | NA | 26.98 (24.41, 30.15) |
| Samoan | NA | NA | NA | 17.27 (15.88, 18.93) | NA |
| Other Pacific Islander | NA | 4.61 (4.35, 4.90) | 6.31 (5.97, 6.69) | 11.83 (11.21, 12.53) | 11.66 (11.06, 12.32) |

**eTable 3. Methamphetamine overdose crude mortality rates per 100,000 among disaggregated Asian American and Native Hawaiian/Pacific Islander populations, 2018 to 2022**

|  | **Crude mortality rate per 100,000 population (95% CI)** | | | | |
| --- | --- | --- | --- | --- | --- |
| **Race** | **2018** | **2019** | **2020** | **2021** | **2022** |
| Asian Americans (aggregated) | 0.98 (0.84, 1.12) | 1.19 (1.04, 1.35) | 2.36 (2.15, 2.57) | 2.52 (2.30, 2.73) | 3.17 (2.93, 3.41) |
| Asian Indian | 1.01 (1, 1.02) | 1.34 (1.33, 1.36) | 1.97 (1.94, 1.99) | 2.45 (2.42, 2.49) | 3.33 (3.29, 3.37) |
| Chinese | 0.45 (0.45, 0.46) | 0.59 (0.58, 0.6) | 0.71 (0.70, 0.71) | 1.10 (1.09, 1.11) | 1.11 (1.09, 1.12) |
| Filipino | 0.62 (0.61, 0.62) | 0.94 (0.92, 0.95) | 2.05 (2.02, 2.09) | 2.09 (2.06, 2.13) | 3.47 (3.41, 3.52) |
| Japanese | NA | 1.72 (1.68, 1.76) | 2.94 (2.87, 3.01) | 2.42 (2.37, 2.49) | 3.76 (3.65, 3.88) |
| Korean | 1.57 (1.53, 1.60) | 2.19 (2.15, 2.23) | 4.82 (4.71, 4.92) | 4.77 (4.67, 4.88) | 5.26 (5.16, 5.37) |
| Vietnamese | 1.13 (1.11, 1.15) | 1.33 (1.31, 1.36) | 3.29 (3.22, 3.36) | 3.59 (3.51, 3.67) | 4.24 (4.15, 4.33) |
| Other Asian | 2.02 (1.99, 2.05) | 1.78 (1.75, 1.81) | 4.50 (4.43, 4.57) | 4.15 (4.09, 4.21) | 4.91 (4.83, 4.99) |
| Native Hawaiians/Pacific Islanders (aggregated) | 1.63 (0.87, 2.78) | 2.85 (1.81, 4.28) | 4.27 (2.99, 5.91) | 8.92 (7.04, 11.15) | 8.19 (6.41, 10.32) |
| Hawaiian | NA | NA | NA | 10.25 (9.71, 10.85) | 5.39 (5.08, 5.75) |
| Guamanian | NA | NA | NA | NA | 26.98 (24.41, 30.15) |
| Samoan | NA | NA | NA | 17.27 (15.88, 18.93) | NA |
| Other Pacific Islander | NA | 4.61 (4.35, 4.90) | 6.31 (5.97, 6.69) | 11.83 (11.21, 12.53) | 11.66 (11.06, 12.32) |
